# Social Determinants of Healthy Aging: An Investigation using the All of Us Cohort

**DOI:** 10.1101/2025.01.02.25319907

**Authors:** Wei-Han Chen, Yao-An Lee, Huilin Tang, Chenyu Li, Ying Lu, Yu Huang, Rui Yin, Melissa J. Armstrong, Yang Yang, Gregor Štiglic, Jiang Bian, Jingchuan Guo

**Affiliations:** Department of Pharmaceutical Outcomes and Policy, University of Florida, Gainesville, Florida; Department of Biomedical Informatics, University of Pittsburgh, Pittsburgh, Pennsylvania; Department of Health Outcomes and Bioinformatics, University of Florida, Gainesville, Florida; Fixel Institute for Neurological Diseases, Department of Neurology, University of Florida, Gainesville, Florida; Databricks Inc.; Faculty of Health Sciences, University of Maribor, Maribor, Slovenia; Usher Institute, University of Edinburgh, Edinburgh, UK

## Abstract

**Introduction:** The increasing aging population raises significant concerns about the ability of individuals to age healthily, avoiding chronic diseases and maintaining cognitive and physical functions. However, the pathways through which SDOH factors are associated with healthy aging remain unclear.

**Methods:** This retrospective cohort study used the registered tier data from the *All of Us* Research Program (AoURP) registered tier dataset v7. Eligible study participants are those aged 50 and older who have responded to any of the SDOH survey questions with available EHR data. Three different algorithms were trained (logistic regression [LR], multi-layer perceptron [MLP], and extreme gradient boosting [XGBoost]). The outcome is healthy aging, which is measured by a composite score of the status for 1) comorbidities, 2) cognitive conditions, and 3) mobility function. We evaluate the model performance by area under the receiver operating characteristic curve (AUROC) and assess the fairness of best-performed model through predictive parity. Feature importance is analyzed using SHapley Additive exPlanations (SHAP) values.

**Results:** Our study included 99,935 participants aged 50 and above, and the mean (SD) age was 74 (9.3), with 55,294 (55.3%) females, 67,457 (67.5%) Whites, 11,109 (11.1%) Hispanic ethnicity, and 44,109 (44.1%) are classified as healthy aging. Most of the individuals lived in their own house (64%), were married (51%), obtained college or advanced degrees (74%), and had Medicare (56.2%). The best predictive model was XGBoost with random oversampler, with a performance of AUROC [95% CI]: 0.793 [0.788-0.796], F1 score: 0.697 [0.692-0.701], recall: 0.739 [0.732-0.748], precision: 0.659 [0.655-0.663], and accuracy: 0.716 [0.712-0.720], and the XGBoost model achieved predictive parity by similar positive and negative predictive values across race and sex groups (0.86-1.06). In feature importance analysis, health insurance type is ranked as the most predictive feature, followed by employment status, substance use, and health insurance coverage (yes/no).

**Conclusion:** In this cohort study, XGBoost model accurately predicted individuals achieving healthy aging, outperforming LR and MLP. Our findings underscore the significant role of health insurance in contributing to healthy aging.

## INTRODUCTION

Aging is an inevitable biological process characterized by a gradual decline in physiological functions, leading to increased vulnerability to diseases and death. The concept of healthy aging has emerged as a critical focus in geriatric research and public health.(1–3) Healthy aging refers to the process of developing and maintaining functional abilities that enable well-being in older age.(4–6) It encompasses physical, mental, cognitive, and social well-being, allowing individuals to live independently and enjoy a good quality of life despite the natural aging process. The increasing proportion of older individuals in the global population has intensified the need to understand and promote healthy aging, making it a vital area of study.(7) In 2020, nearly 1 in 6 Americans were 65 years or older, and this group is estimated to constitute 23% of the total US population in 2050.(8,9)

Social determinants of health (SDOH) — the conditions where people are born, grow, work, live, and age — play a crucial role in individuals’ health, influencing the aging process and the ability to age healthily.(10) SDOH includes factors such as socioeconomic status, education, neighborhood and physical environment, employment, social support networks, and access to healthcare.(11–13) Previous studies have demonstrated that these SDOH can significantly affect an individual’s health outcomes by influencing behaviors, exposures, and access to resources necessary for maintaining health.(14–18) Individuals with higher socioeconomic status, better education, and stronger social support tend to have better health outcomes and a higher likelihood of healthy aging.(19–21) Addressing disparities in SDOH is therefore essential for promoting health equity and improving the quality of life for older adults, especially those socioeconomically disadvantaged groups.(22–24)

Existing studies on the impact of SDOH on healthy aging are limited.(25) For instance, Sowa et al. have identified a set of predictors using health surveys in Europe, however, they focused only on lifestyle and psychosocial factors and did not consider many other SDOH.(26) On the other hand, the application of machine learning (ML) models has shown great promise in predicting health outcomes.(27) Other studies that applied ML techniques have mainly focused on biological or physiological factors in healthy aging,(28) none have studied SDOH.

To fill the gap, the objective of this study is to develop a prediction model of healthy aging by leveraging a large cohort of older adults from the AoU and advanced ML techniques. Understanding the relationship between SDOH and healthy aging holds significant clinical and policy implications. Clinically, this knowledge enables healthcare providers to create more personalized care plans that address both medical and social factors influencing a patient’s health. On the policy side, identifying key SDOH linked to healthy aging can guide targeted interventions and resource allocation, fostering public health strategies that promote healthy aging across diverse populations. Additionally, we also evaluated the fairness of the ML models in predicting healthy aging, ensuring that they do not perpetuate existing disparities and can be applied equitably across different demographic groups. Lastly, we identified the top predictors for healthy aging using SHapley Additive exPlanations (SHAP) values, a well-established explainable ML method, which could inform the development of targeted interventions and policies to support healthy aging by addressing the most influential SDOH.

## METHODS

### Data Source and Study Population

We used the registered tier data from the *All of Us* (AoU) Research Program registered tier dataset v7.(29) The AoU was a nationwide program funded by the National Institute of Health, which aimed to provide diverse and comprehensive information among under-represented groups. The database included survey questions (e.g., lifestyle, demographic, and social determinants of health) and electronic health records (EHR).(30) Both survey questions and EHR were standardized and could be mapped utilizing Observational Medical Outcomes Partnership (OMOP) Common Data Model infrastructure.(31) We included individuals aged ≥50 years of age who have responded to any of the SDOH survey questions with available EHR data.

### Study Outcome

The primary outcome is a dichotomous score of healthy aging, which was measured by a composite score of the status for 1) comorbidities, 2) cognitive conditions, and 3) mobility function. Charlson comorbidity index (CCI) by Quan. et al(32) was used for assessing comorbidity status. We modified the original CCI algorithm to exclude age as a parameter (referred to as modified CCI [mCCI]) since our goal was to predict healthy aging. Secondly, we assessed the cognitive conditions by ICD-9 and −10 CM codes with a diagnosis of mild cognitive impairment (MCI). Lastly, to assess the mobility function, we identified individuals in assisted living using CPT/HCPCS codes and records of discharge locations. An individual aged over 75 is classified as experiencing healthy aging if they have a composite score of 0, which includes an mCCI score of 0, no MCI, and are not in assisted living. A composite score greater than 0 indicates non-healthy aging.

We also defined a secondary cohort as a composite score of 0, with age greater than 85 classified as healthy aging, otherwise as non-healthy aging. Two distinct cohorts were then created for primary outcome and the secondary outcome analysis, respectively. The secondary cohort analysis allows us to examine whether the association between SDOH and healthy aging hold consistent when applying a more stringent definition of healthy aging. Consistent results across both cohorts would reinforce the robustness of our findings across varying definitions of healthy aging.

### Study Design

We adopted a retrospective cohort study design and illustrated the cohort selection process in **Figure 1**. Patients aged under 75 with an mCCI score of 0 are excluded from the analysis in primary cohort. For the secondary cohort, this exclusion extends to patients aged under 85 with an mCCI score of 0.

**Figure 1.**
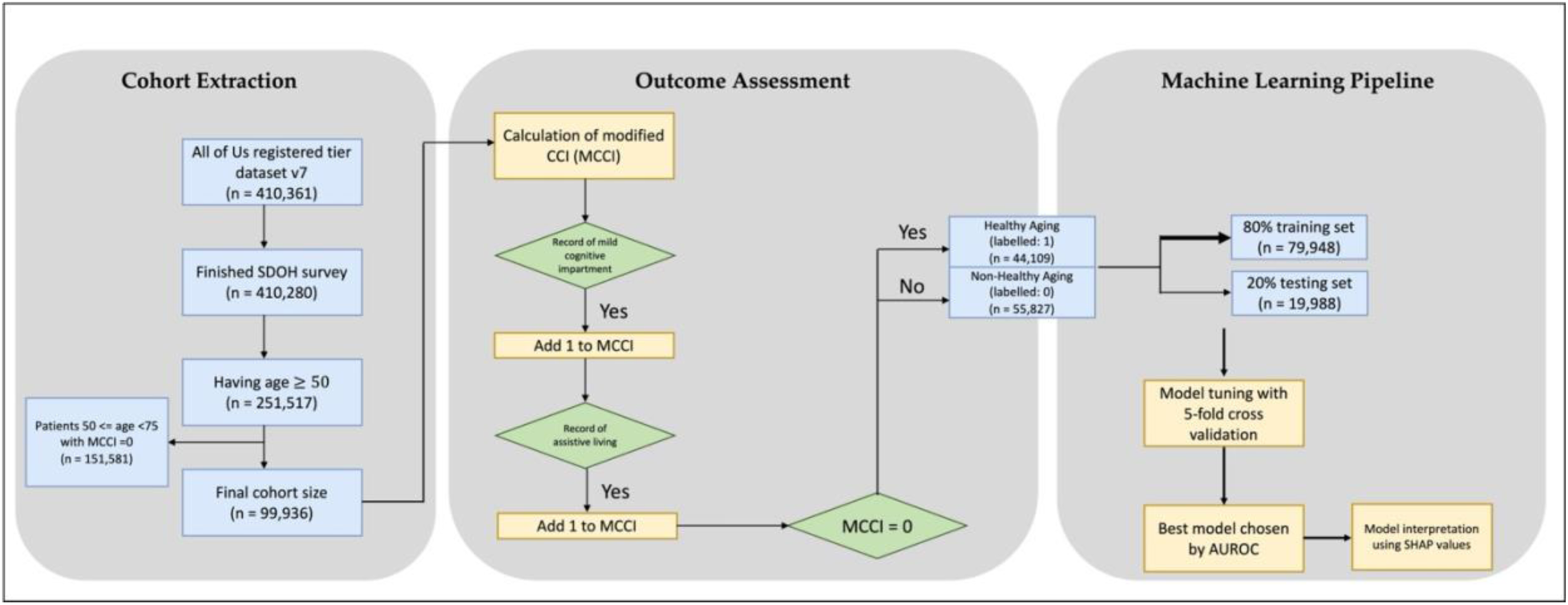
The overall workflow including participant selection, outcome assessment, and machine learning pipeline.

### Potential Risk Factors

Potential risk factors (i.e., input features) were SDOH information collected from multifaceted survey questions, including The Basics (demographic information), Lifestyle (smoking, alcohol use, substance use, etc.), Healthcare Access & Utilization (access to and use of health care resources), and Social Factors (neighborhood, social life, stress, etc.). Self-reported race and gender were also recorded and included in the analysis. We reported the counts (percentages) for categorical variables and median (interquartile range) for continuous variables.

### Statistical analysis

We aimed to use SDOH features to develop a machine learning model to predict healthy aging. Three machine learning algorithms were applied: logistic regression (LR), multi-layer perceptron (MLP), and Extreme Gradient Boosting(33) (XGBoost).

Regularization was employed in both logistic regression (lasso [L1](34), ridge [L2](35), and ElasticNet(36)) and XGBoost (alpha [L1] and lambda [L2]) to reduce overfitting. Following machine learning best practices, we split the data into training and testing with a ratio of 8:2. To account for target class imbalance, we employed both random over-sampling and random under-sampling methods and compared their performance for further analyses.(37) For random over-sampling, we increased the minority class to match the size of the majority class, resulting in a final balanced distribution of 50% for each class. Similarly, for random under-sampling, we reduced the majority class to match the size of the minority class, also achieving a balanced class distribution.

After hyperparameters tuning using Bayesian optimization with 5-fold cross-validation over 100 iterations to optimize the area under the receiver operating characteristic curve (AUROC), we reported the performance metrics of the testing set including AUROC, precision, recall, F1 score, and specificity. In addition, we obtained the 95% confidence intervals (CI) of the performance metrics by bootstrap method with 50 iterations. The best model is selected based on AUROC and the potential clinical application with the goal of a higher F1 score, showing the balance between precision and recall.

We then assessed the fairness of the machine learning model selected by comparing the ratios of metrics such as positive predicted value (PPV), negative predicted value (NPV), false positive rate (FPR), true positive rate (TPR), false negative rate (FNR), and overall accuracy across race and gender. We designated non-Hispanic Whites and females as the privileged groups for race and gender, respectively, and identified Black and males as the protected groups for these categories. Lastly, we adopted SHapley Additive exPlanations (SHAP) values to identify and rank the most important features, with a view to providing explainability and improved clinical decision-making.(38)

All analyses were performed in Python (version 3.10 with libraries such as Scikit-learn, Imbalanced-learn). The study followed the STROBE cohort reporting guideline.(39)

## RESULTS

### Descriptive Statistics

In the primary cohort, 99,936 eligible older adults aged 50 or older who had responded to SDOH survey questions were included, and 44,109 (44%) were identified as healthy aging (age ≥75 and a composite condition score of 0, **Table 1**). The mean (SD) age was 74 (9.3) years, with 55,294 (55.3%) females, 41,977 (42.0%) males. Of the cohort, 67,457 (67.5%) were White, 14,612 (14.6%) were Black or African American, 11,109 (11.1%) having Hispanic ethnicity. The median (IQR) of the mCCI was 1 (0–2). Most of the individuals lived in their own house (64%), were married (51%), obtained college or advanced degrees (74%), and had Medicare (56.2%).

**Table 1.** Descriptive analysis of participants demographic information.

| Variables | Primary Cohort, No. (%) |  |  |  | Secondary Cohort, No. (%) |  |  |  |
| --- | --- | --- | --- | --- | --- | --- | --- | --- |
|  | Overall,<br>n =<br>99,936 | Healthy<br>aging (75),<br>n = 44,109<br>(44.1%) | Non-healthy<br>aging, n =<br>55,827<br>(55.9%) | P | Overall,<br>n =<br>62,475 | Healthy<br>aging (85),<br>n = 6,648<br>(10.6%) | Non-healthy<br>aging, n =<br>55,827<br>(89.4%) | P |
| **Age, mean (SD)** | 74.0<br>(9.3) | 79.9 (4.0) | 69.4 (9.6) | <.001 | 71.2<br>(10.6) | 87.2 (1.8) | 69.4 (9.6) | <.001 |
| **Sex** |  |  |  | <.001 |  |  |  | <.001 |
| Male | 41,977<br>(42.0%) | 20,493<br>(46.5%) | 21,484<br>(38.5%) |  | 24,671<br>(39.5%) | 3,187<br>(47.9%) | 32,839<br>(58.8%) |  |
| Female | 55,294<br>(55.3%) | 22,455<br>(50.9%) | 32,839<br>(58.8%) |  | 36,101<br>(57.8%) | 3,262<br>(49.1%) | 21,484<br>(38.5%) |  |
| Other | 2,665<br>(2.7%) | 1,161 (2.6%) | 1,504<br>(2.7%) |  | 1,703<br>(2.7%) | 199 (3.0%) | 1,504<br>(2.7%) |  |
| **Race** |  |  |  | <.001 |  |  |  | <.001 |
| White | 67,457<br>(67.5%) | 33,916<br>(76.9%) | 33,541<br>(60.1%) |  | 38,802<br>(62.1%) | 5,261<br>(79.1%) | 33,541<br>(60.1%) |  |
| Black or African American | 14,612<br>(14.6%) | 3,597 (8.2%) | 11,015<br>(19.7%) |  | 11,437<br>(18.3%) | 422 (6.4%) | 11,015<br>(19.7%) |  |
| Asian | 1,775<br>(1.8%) | 941 (2.1%) | 834 (1.5%) |  | 954<br>(1.5%) | 120 (1.8%) | 834 (1.5%) |  |
| Other/Unknown | 16,092<br>(16.1%) | 5,655<br>(12.8%) | 10,437<br>(18.7%) |  | 11,282<br>(18.1%) | 845 (12.7%) | 10,437<br>(18.7%) |  |
| **Modified Charlson comorbidity index, median (IQR)** | 1 (0-2) | 0 (0-0) | 2 (1-3) | <.001 | 2 (1-3) | 0 (0-0) | 2 (1-3) | <.001 |
| **Ethnicity** |  |  |  | <.001 |  |  |  | <.001 |
| Not Hispanic or Latino | 84,332<br>(84.4%) | 38,693<br>(87.7%) | 45,639<br>(81.8%) |  | 51,477<br>(82.4%) | 5,838<br>(87.8%) | 45,639<br>(81.8%) |  |
| Hispanic or Latino | 11,109<br>(11.1%) | 3,295 (7.5%) | 7,814<br>(14.0%) |  | 8,270<br>(13.2%) | 456 (6.9%) | 7,814<br>(14.0%) |  |
| Other/Unknown | 4,495<br>(4.5%) | 2,121 (4.8%) | 2,374<br>(4.2%) |  | 2,728<br>(4.4%) | 354 (5.3%) | 2,374<br>(4.2%) |  |
| **Housing Status** |  |  |  | <.001 |  |  |  | <.001 |
| Own | 63,907<br>(63.9%) | 32,924<br>(74.6%) | 30,983<br>(55.5%) |  | 35,729<br>(57.2%) | 4,746<br>(71.4%) | 30,983<br>(55.5%) |  |
| Rent | 27,278<br>(27.3%) | 8,018<br>(18.2%) | 19,260<br>(34.5%) |  | 20,547<br>(32.9%) | 1,287<br>(19.4%) | 19,260<br>(34.5%) |  |
| Other/Unknown | 8,751<br>(8.8%) | 3,167 (7.2%) | 5,584<br>(10.0%) |  | 6,199<br>(9.9%) | 615 (9.2%) | 5,584<br>(10.0%) |  |
| **Current Marital Status** |  |  |  | <.001 |  |  |  | <.001 |
| Married | 51,179<br>(51.2%) | 25,234<br>(57.2%) | 25,945<br>(46.5%) |  | 29,294<br>(46.9%) | 3,349<br>(50.4%) | 25,945<br>(46.5%) |  |
| Divorced | 17,556<br>(17.6%) | 6,900<br>(15.6%) | 10,656<br>(19.1%) |  | 11,503<br>(18.4%) | 847 (12.7%) | 10,656<br>(19.1%) |  |
| Widowed | 11,711<br>(11.7%) | 6,626<br>(15.0%) | 5,085<br>(9.1%) |  | 6,864<br>(11.0%) | 1,779<br>(26.8%) | 5,085<br>(9.1%) |  |
| Never married | 11,226<br>(11.2%) | 2,690 (6.1%) | 8,536<br>(15.3%) |  | 8,824<br>(14.1%) | 288 (4.3%) | 8,536<br>(15.3%) |  |
| Other/Unknown | 8,264<br>(8.3%) | 2,659 (6.0%) | 5,605<br>(10.0%) |  | 5,990<br>(9.6%) | 385 (5.8%) | 5,605<br>(10.0%) |  |
| **Education Level** |  |  |  | <.001 |  |  |  | <.001 |
| College, graduate or advanced degree | 74,320<br>(74.4%) | 35,657<br>(80.8%) | 38,663<br>(69.3%) |  | 43,773<br>(70.0%) | 5,110<br>(76.9%) | 9,948<br>(17.8%) |  |
| High school or GED | 14,895<br>(14.9%) | 4,947<br>(11.2%) | 9,948<br>(17.8%) |  | 10,864<br>(17.4%) | 916 (13.8%) | 38,663<br>(69.3%) |  |
| Less than high school | 7,403<br>(7.4%) | 2,194 (5.0%) | 5,209<br>(9.3%) |  | 5,610<br>(9.0%) | 401 (6.0%) | 5,209<br>(9.3%) |  |
| Other/Unknown | 3,318<br>(3.3%) | 1,134 (3.0%) | 2,007<br>(3.6%) |  | 2,228<br>(3.6%) | 221 (3.3%) | 2,007<br>(3.6%) |  |
| **Health Insurance\*** |  |  |  |  |  |  |  | * |
| Medicare | 56,150<br>(56.2%) | 32,399<br>(73.5%) | 23,751<br>(42.5%) |  | 28,525<br>(45.7%) | 4,774<br>(71.8%) | 23,751<br>(42.5%) |  |
| Medicaid | 14,553<br>(14.6%) | 2,886 (6.5%) | 11,667<br>(20.9%) |  | 12,082<br>(19.3%) | 415 (6.2%) | 11,667<br>(20.9%) |  |
| Private/Other | 48,914<br>(49%) | 22,482 (51%) | 26,432<br>(47.3%) |  | 29,936<br>(48%) | 3,504<br>(52.7%) | 26,432<br>(47.3%) |  |
| None | 143<br>(0.1%) | 32 (0.1%) | 111 (0.2%) |  | 115<br>(0.2%) | <20 | 111 (0.2%) |  |
| Unknown | 1,073<br>(1.1%) | 379 (0.9%) | 694 (1.2%) |  | 768<br>(1.2%) | 74 (1.1%) | 694 (1.2%) |  |
| **Substance use\*** |  |  |  |  |  |  |  | * |
| Marijuana | 40,084<br>(40.1%) | 15,215<br>(34.5%) | 24,869<br>(44.5%) |  | 25,970<br>(41.6%) | 1,101<br>(16.6%) | 24,869<br>(44.5%) |  |
| Cocaine | 13,417<br>(13.4%) | 3,383 (7.7%) | 10,034<br>(18%) |  | 10,181<br>(16.3%) | 147 (2.2%) | 10,034<br>(18.0%) |  |
| Opioids | 8,414<br>(8.4%) | 2,380 (5.4%) | 6,034<br>(10.8%) |  | 6,257<br>(10%) | 223 (3.3%) | 6,034<br>(10.8%) |  |
| Other | 29,268<br>(29.3%) | 10,306<br>(23.4%) | 18,962<br>(34%) |  | 19,736<br>(31.6%) | 774 (11.6%) | 18,962<br>(34%) |  |
| None | 45,982<br>(46%) | 22,171<br>(50.3%) | 23,811<br>(42.7%) |  | 28,180<br>(45.1%) | 4,369<br>(65.7%) | 23,811<br>(42.7%) |  |
| Unknown/skipped | 7,799<br>(7.8%) | 4,180 (9.5%) | 3,619<br>(6.5%) |  | 4,355<br>(7%) | 736 (11.1%) | 3,619<br>(6.5%) |  |

\*Allow having multiple responses per participant

In the secondary cohort, 62,475 participants were included, and 6,648 (10.6%) were identified as healthy aging (i.e., age ≥85 and a composite condition score of 0, **Table 1**). The mean (SD) age is 71 (10.6) years, with 36,101 (58%) females, 24,671 (40%) males. 38,802 (62%) were White, 11,437 (18%) were Black or African American, 8,270 (13%) having Hispanic ethnicity. The median (IQR) of the mCCI was 2 (1–3). Similarly, most of the individuals lived in their own house (57%), were married (46.9%), obtained college or advanced degrees (70%), and had Medicare (46%).

### Model Performance and Selection

Performance metrics on the test dataset and the AUROC for the three models are presented in **Figure 2 and Supplemental table S1.** Bootstrapped performance with 95% CI over 50 iterations for the best algorithm are included in **Supplemental table S2.** Overall, all three models achieved decent prediction performance with AUROC >0.7.

**Figure 2.**
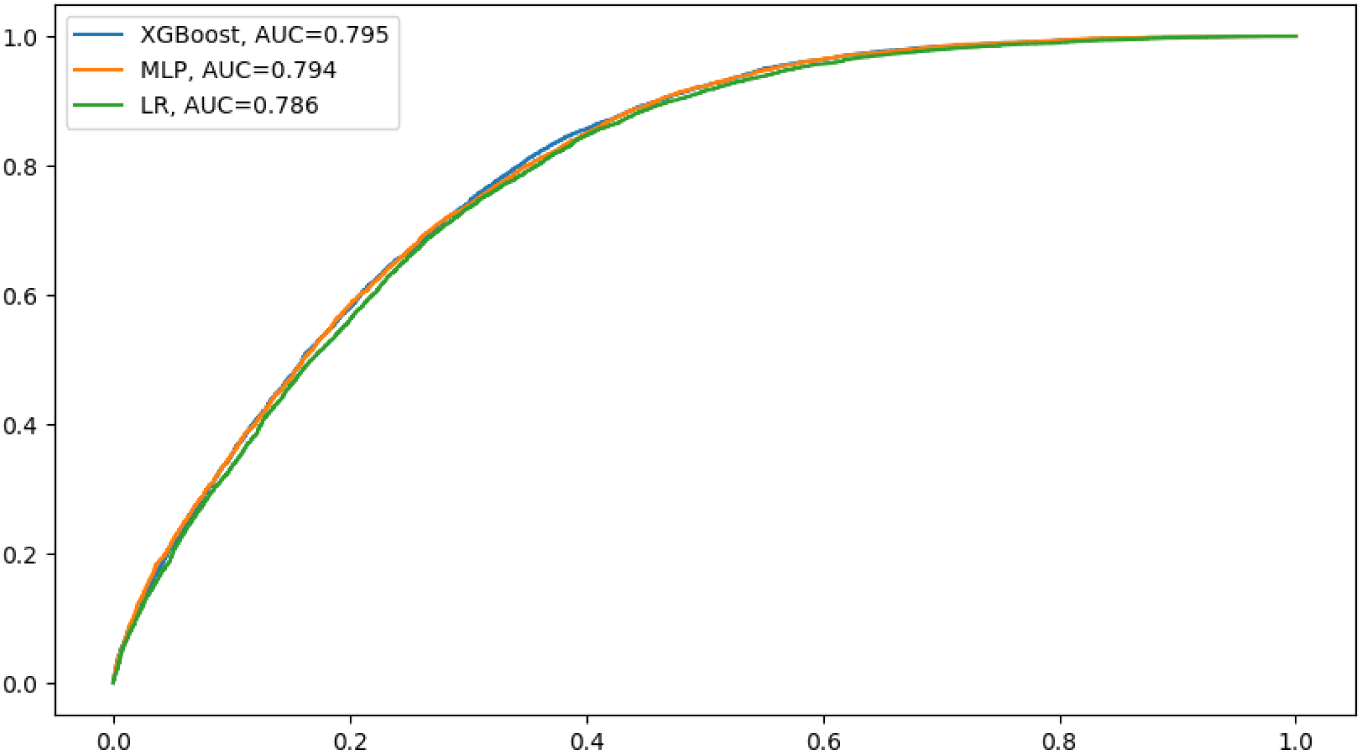
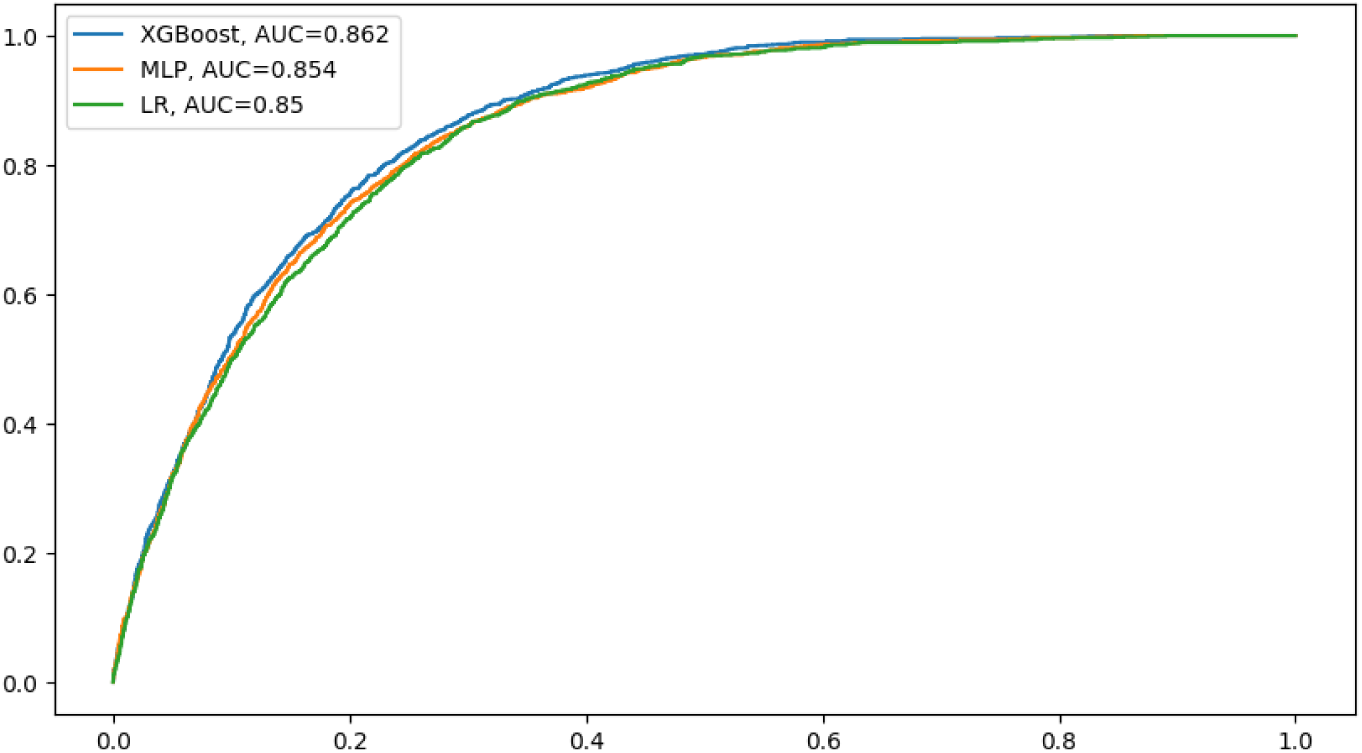
Comparison of model performance on test datasets with area under the receiver operating characteristic curve. *XGBoost: extreme gradient boosting, LR: logistic regression, MLP: multilayer perceptron.

Among them, we found that the XGBoost model with over-sampling adjustments (AUROC: 0.795 and 0.862 for primary and secondary cohort, respectively) shows superior performance. This outperformed both the LR model (AUROC: 0.786 and 0.85) and the MLP model (AUROC: 0.794 and 0.854). Though the AUROC was comparable between XGBoost and MLP, the F1 score and other metrics of MLP classifier are much lower than those of XGBoost classifier. Detailed machine learning algorithm and tuning values are included in **Supplemental table S3**.

### Model Fairness Assessment

**Table 2** demonstrates the fairness metrics across gender and race for the best selected model, XGBoost. The ratios of accuracies and PPVs showed no evidence of model biases towards a specific population.

**Table 2.**
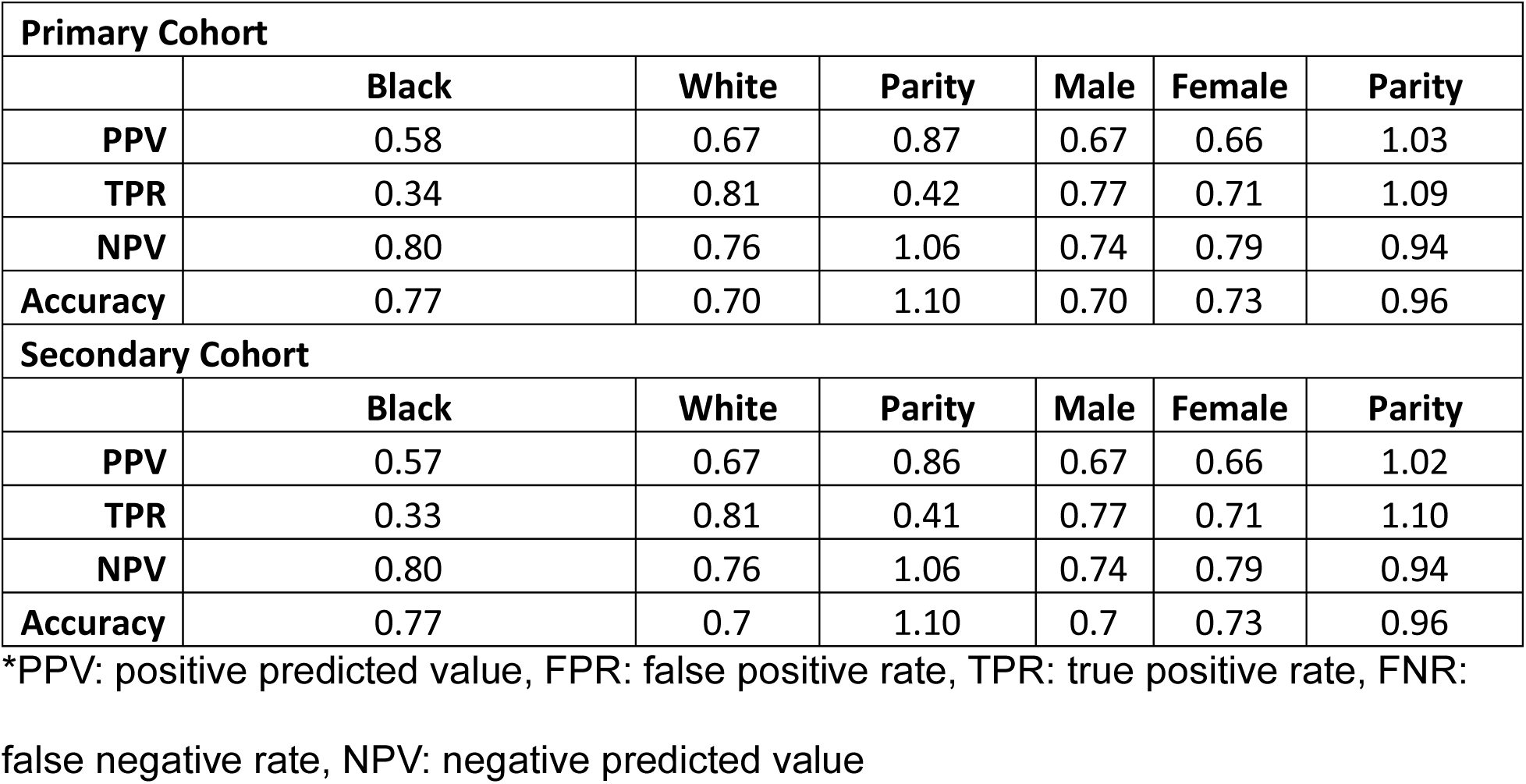
Fairness metrics for XGBoost across gender and race.

### Feature Importance Analysis

**Figure 3** shows the SHAP values to explain the healthy aging prediction of XGBoost model (best performance). In both cohorts, health insurance type (e.g., Medicare, Medicaid, insurance purchased from a company) is ranked as the most predictive feature (SHAP value: 0.595), followed by employment status (0.233), substance use (0.171), health insurance coverage (yes/no, SHAP value 0.143). The direction of the plots revealed that all top 10 features were positively (red on the right in figure 3) associated with healthy aging.

**Figure 3.**
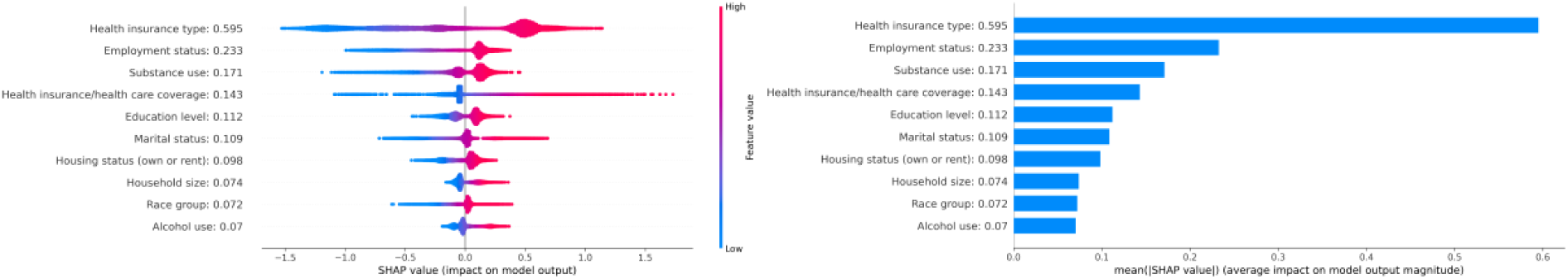
Distribution of the SHAP values for the top 10 features based on the highest mean absolute SHAP value (left panels) and their mean absolute contribution of the top 10 features, ranked by their average SHAP value (right panels). Each test sample is depicted as a point for every feature, with the x-axis indicating whether the feature’s effect on the model’s prediction is positive (red on the right) or negative (blue on the right). The color of each point reflects the feature’s value, and this color scale is adjusted individually according to the value range present in the dataset.

**Figure 4.**
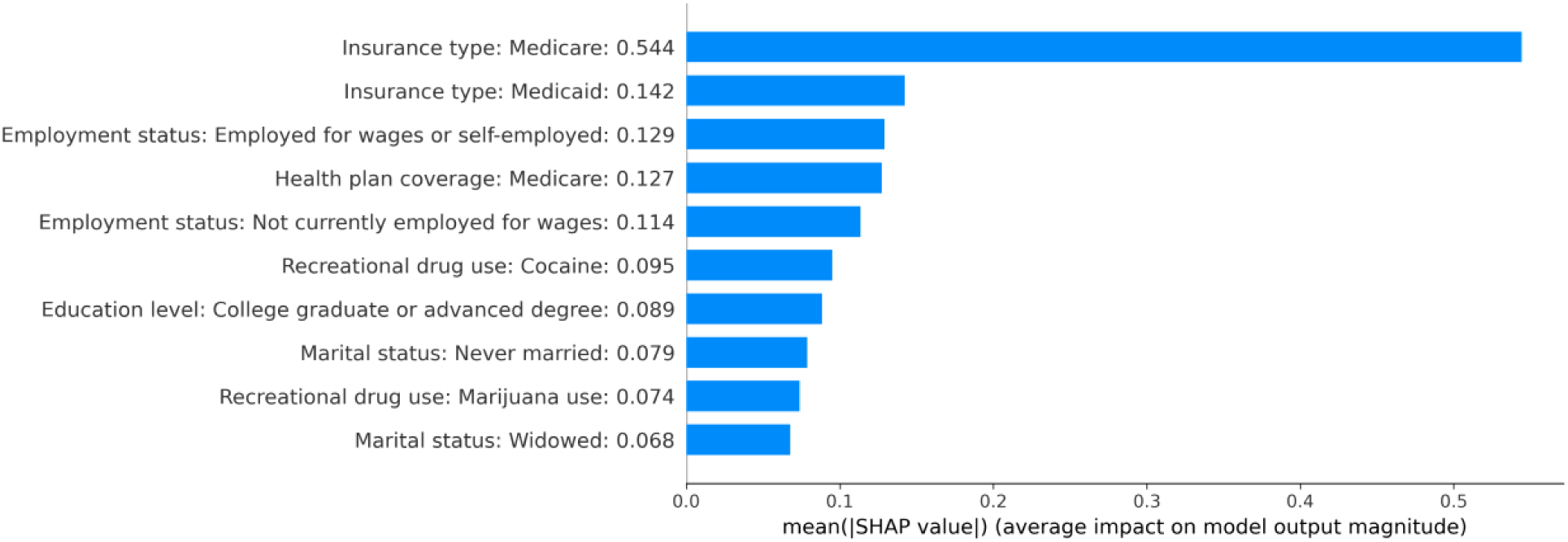
Mean absolute contribution of the top 10 features, ranked by their average SHAP value.

## DISCUSSION

This cohort study leverages the AoU datasets, which not only included diverse populations from historically underrepresented groups and racial/ethnic minority groups, but also provided a rich source of SDOH through standardized OMOP data infrastructure. Our findings suggest that machine learning models could accurately predict healthy aging using SDOH information and highlight the potential for integrating SDOH factors in clinical decision-making to enhance predictive accuracy.

It is noteworthy that our work included high-dimensional SDOH with large-scale population and an explainable ML framework. A few of previous studies were available to include SDOH across several domains, such as neighborhood environment, education access, etc..(40,41) This study also added values by the integration of objective measures from EHRs with detailed survey data on SDOH, providing a more comprehensive assessment of healthy aging compared to studies relying solely on self-reported data.(26)

Our models showed fairness in predictive parity, where the ratios of both positive (PPV) and negative predictive values (NPV) are close to 1. Ensuring that ML models do not discriminate against different racial or ethnic groups is crucial, as these models must perform equitably independent on sensitive features. In our context, if healthy aging is less accurately identified in disadvantaged groups, it may lead to unnecessary and potentially harmful treatments, thereby increasing their financial burden and causing undue harm. Therefore, maintaining equal PPV and NPV across different demographic groups is imperative to prevent such disparities and ensure equitable and healthcare outcomes.(42)

Some machine learning classifiers are notorious as a “black box” where excellent performance is often obtained at the cost of lacking interpretability.(43–47) In the feature importance analysis, health insurance was the strongest positive SDOH factor for predicting healthy aging. Our study identified top SDOH factors from several domains positively associated with healthy aging: health insurance type, employment status, education level, marital status, housing status. These aligned with previous studies indicating that higher socioeconomic status, including higher income and education level, was associated with better health outcomes.(28,48) Although the precise mechanism of marital and housing statuses on healthy aging has not yet been identified, studies have shown that there was an intricate pattern associated with mental health and chronic diseases.(49,50) Our study added the evidence that they could also be related to healthy aging.

Past research has suggested that substance use and related drug overdoses may have contributed to lower life expectancy,(51–53) however, we found that substance and alcohol use were positively associated with healthy aging. We suggest the following probable underlying causes of the results. First, it is possible that substance and alcohol users in our cohort be healthier than those in the general population from previous studies since our participants mainly participated AoU voluntarily rather than being randomly recruited into the program. Secondly, some substance and alcohol users may have altered their health behaviors following enrollment in the program, namely, Hawthorne effect, where individuals knowingly adopt a healthier behavior when they were being assessed in a research program.(54,55) Thus, there may be difference in substance and alcohol use status between the baseline period and the follow-up period. Lastly, according to statistics from Centers for Disease Control and Prevention, the drug overdose death rates were higher among groups aged 25-44 (∼50 deaths per 100,000 population) compared to those aged over 55 (5-35 death per 100,000 population).(56) Thus, our results could be affected by selection bias since we only included participants aged over 50 in our study, whose health behaviors are general substance and alcohol use populations. These factors could contribute to the unexpected positive association between substance use and healthy aging in our study.

This study has some limitations. First, to date, a unanimous definition of healthy aging has not yet been reached.(57) Our definition of healthy aging, while based on objective measures, may not capture all constructs of ‘healthy aging’ such as quality of life, social engagement, and subjective well-being. Also, CCI limited the spectrum of comorbidities. For instance, some may not consider Parkinson’s disease as healthy aging, however, it is not covered in CCI.

Secondly, while the All of Us Research Program provides a large and diverse dataset, it may not be fully representative of the U.S. population. Participants in All of Us are volunteers who agreed to share their health data, which could introduce selection bias. These individuals may be more health-conscious, have better access to healthcare, or obtained higher educational degrees than the general population, potentially leading to an overestimation of healthy aging in our sample. While the generalizability of our findings is limited to the participants in AoU, it is important to note that the cohort is wide across the nation. Furthermore, the representation of racial and ethnic minority groups has improved in AoU, which enhances the applicability of our results to a more diverse population.(58) However, caution should still be exercised when extrapolating these findings to other populations or other clinical settings.

Thirdly, while we attempted to account for a wide range of SDOH factors, there may still be unmeasured confounders. For instance, we did not have data on lifelong health behaviors or early-life exposures that could significantly impact aging trajectories.

## CONCLUSION

In this cohort study utilizing the AoU database, our machine learning model effectively predicted individuals likely to achieve healthy aging, emphasizing the critical influence of health insurance on this outcome. The findings highlight that access to health insurance is not merely a facilitator of healthcare services but a pivotal determinant of long-term health outcomes in older adults. By addressing the gaps in health insurance, policymakers can contribute to the promotion of healthy aging across diverse populations, ultimately leading to improved quality of life. The integration of health insurance into public health strategies could therefore be a powerful tool in enhancing the overall well-being of aging populations.

## Data Availability

This dataset for this study is sourced from the All of Us Research Program Registered tier dataset v7, which is not publicly available. The authors have obtained all necessary permissions to access and utilize the All of Us Research Program registered tier dataset for this study. Researchers who are interested in accessing the All of Us Research Program dataset can apply for access through the All of Us Research Hub. For more information and to initiate the application process, please visit the All of Us Research Hub website at https://www.researchallofus.org/.

