## supplemental table 1-3 for "Social Determinants of Healthy Aging: An Investigation using the All of Us Cohort"

### S1. Performance metrics for the three algorithms

| S1. Performance metrics of the testing datasets | | | | | | |
|---|---|---|---|---|---|---|
| **Primary Cohort** | **Under-sampling** | | | **Over-sampling** | | |
| | **LR** | **XGB** | **MLP** | **LR** | **XGB** | **MLP** |
| AUROC | 0.786 | 0.796 | 0.790 | 0.786 | 0.795 | 0.794 |
| F1 score | 0.704 | 0.716 | 0.716 | 0.702 | 0.717 | 0.717 |
| Recall | 0.766 | 0.806 | 0.822 | 0.765 | 0.803 | 0.814 |
| Precision | 0.650 | 0.645 | 0.634 | 0.649 | 0.648 | 0.640 |
| Accuracy | 0.715 | 0.718 | 0.712 | 0.713 | 0.720 | 0.716 |
| **Secondary Cohort** | **Under-sampling** | | | **Over-sampling** | | |
| | **LR** | **XGB** | **MLP** | **LR** | **XGB** | **MLP** |
| AUROC | 0.850 | 0.796 | 0.854 | 0.857 | 0.795 | 0.886 |
| F1 score | 0.411 | 0.716 | 0.408 | 0.418 | 0.717 | 0.442 |
| Recall | 0.807 | 0.806 | 0.826 | 0.802 | 0.803 | 0.869 |
| Precision | 0.276 | 0.645 | 0.271 | 0.283 | 0.648 | 0.297 |
| Accuracy | 0.754 | 0.718 | 0.745 | 0.762 | 0.720 | 0.767 |

### S2. Bootstrapped performance on the test dataset over 50 iterations

| | AUROC (95% CI) | AUPRC (95% CI) | F1 score (95% CI) | Recall (95% CI) | Precision (95% CI) | Accuracy (95% CI) |
|---|---|---|---|---|---|---|
| **Primary Cohort** | | | | | | |
| XGBoost with RUS | 0.792 (0.788-0.796) | 0.707 (0.700-0.713) | 0.697 (0.692-0.701) | 0.742 (0.733-0.749) | 0.658 (0.654-0.662) | 0.716 (0.711-0.719) |
| XGBoost with ROS | 0.793 (0.788-0.796) | 0.708 (0.700-0.714) | 0.697 (0.692-0.701) | 0.739 (0.732-0.748) | 0.659 (0.655-0.663) | 0.716 (0.712-0.720) |
| LR with RUS | 0.785 (0.781-0.789) | 0.697 (0.691-0.702) | 0.703 (0.700-0.707) | 0.772 (0.766-0.778) | 0.646 (0.642-0.652) | 0.713 (0.709-0.717) |
| LR with ROS | 0.785 (0.781-0.789) | 0.698 (0.692-0.703) | 0.679 (0.675-0.685) | 0.696 (0.691-0.703) | 0.663 (0.657-0.668) | 0.710 (0.705-0.714) |
| MLP with RUS | 0.791 (0.787-0.795) | 0.708 (0.702-0.714) | 0.704 (0.693-0.715) | 0.767 (0.729-0.798) | 0.651 (0.641-0.662) | 0.716 (0.711-0.720) |
| MLP with ROS | 0.793 (0.789-0.796) | 0.710 (0.705-0.716) | 0.699 (0.689-0.709) | 0.745 (0.716-0.775) | 0.658 (0.650-0.666) | 0.717 (0.712-0.721) |
| **Secondary Cohort** | | | | | | |

| | | | | | | |
|---|---|---|---|---|---|---|
| XGBoost with RUS | 0.793 (0.789-0.795) | 0.707 (0.702-0.710) | 0.697 (0.693-0.700) | 0.741 (0.733-0.746) | 0.658 (0.655-0.660) | 0.716 (0.714-0.718) |
| XGBoost with ROS | 0.793 (0.788-0.796) | 0.708 (0.700-0.714) | 0.697 (0.692-0.701) | 0.739 (0.732-0.748) | 0.659 (0.655-0.663) | 0.716 (0.712-0.720) |
| LR with RUS | 0.857 (0.854-0.862) | 0.389 (0.379-0.398) | 0.238 (0.235-0.240) | 0.154 (0.151-0.156) | 0.518 (0.510-0.524) | 0.895 (0.894-0.895) |
| LR with ROS | 0.857 (0.854-0.862) | 0.389 (0.379-0.398) | 0.238 (0.235-0.240) | 0.154 (0.151-0.156) | 0.518 (0.510-0.524) | 0.895 (0.894-0.895) |
| MLP with RUS | 0.855 (0.848-0.863) | 0.387 (0.363-0.406) | 0.099 (0.050-0.143) | 0.054 (0.026-0.082) | 0.599 (0.493-0.731) | 0.895 (0.893-0.897) |
| MLP with ROS | 0.860 (0.854-0.868) | 0.397 (0.373-0.417) | 0.138 (0.116-0.161) | 0.079 (0.065-0.095) | 0.574 (0.514-0.648) | 0.896 (0.894-0.898) |

*AUROC: Area under the Receiver Operating Characteristic Curve, AUPRC: Area Under the Precision-Recall Curve, XGBoost: extreme gradient boosting, LR: logistic regression, MLP: multilayer perception, RUS: random undersampling, ROS: random oversampling

S3. Selection of model hyperparameters using Bayesian optimization with 5-fold cross-validation

| **Hyperparameter** | **Tuning range for Bayesian optimization** | **Optimal value chosen** |
|---|---|---|
| **Primary cohort** | | |
| Column sample by tree | Real (0.5, 1, ‘log-uniform’) | 0.5 |
| Learning rate | Real (0.1, 0.5, ‘log-uniform’) | 0.1 |
| Gamma | Real (1e-1, 1, ‘log-uniform’) | 0.14924415627138107 |
| Maximum tree depth | Integer (3, 10) | 9 |
| Minimum child weight | Integer (1, 20) | 8 |
| Number of estimators | Integer (10, 500) | 419 |
| Alpha | Real (1e-5, 100, ‘log-uniform’) | 7.80142448939489 |
| Lambda | Real (1e-5, 100, ‘log-uniform’) | 100.0 |
| Subsample | Real (0.1, 1, ‘log-uniform’) | 1.0 |
| **Secondary cohort** | | |
| Column sample by tree | Real (0.5, 1, ‘log-uniform’) | 0.5236057037849238 |
| Learning rate | Real (0.1, 0.5, ‘log-uniform’) | 0.1 |
| Gamma | Real (1e-1, 1, ‘log-uniform’) | 0.5904806515456753 |
| Maximum tree depth | Integer (3, 10) | 10 |

| | | |
|---|---|---|
| Minimum child weight | Integer (1, 20) | 10 |
| Number of estimators | Integer (10, 500) | 429 |
| Alpha | Real (1e-5, 100, 'log-uniform') | 0.06936620412496958 |
| Lambda | Real (1e-5, 100, 'log-uniform') | 49.25072571620478 |
| Subsample | Real (1e-5, 100, 'log-uniform') | 1.0 |
